# Concurrent longitudinal changes in plasma p-tau217, tau PET, and cognition in preclinical Alzheimer’s disease

**DOI:** 10.1101/2025.03.25.25324632

**Authors:** Philip S. Insel, Niklas Mattsson-Carlgren, Oliver Langford, Vincent M. Caruso, Antoine Leuzy, Christina B. Young, Adam Boxer, Paul S. Aisen, Reisa A. Sperling, Elizabeth C. Mormino, Michael C. Donohue

**Author notes:** Corresponding author: Philip S. Insel, Department of Psychiatry and Behavioral Sciences, University of California, San Francisco, CA, USA.

## Abstract

**Background:** Plasma p-tau217 and tau PET are biomarkers reflecting partly different aspects of Alzheimer’s disease pathology. Plasma p-tau217 may become abnormal earlier in the disease process, potentially capturing the initial alterations of tau metabolism, whereas tau PET provides spatially detailed information about aggregated tau that could more closely reflect ongoing changes impacting cognitive outcomes. How these two markers evolve over time, whether they diverge in their relationship to cognition, and their potential to assess disease modification in treatment trials are unknown.

**Methods:** Analyses included data from 1707 participants (1169 Aβ+, 538 Aβ–) enrolled in the A4 and LEARN studies, with a subset of 443 (388 Aβ+, 55 Aβ–) receiving tau PET scans. All participants were aged 65–85, cognitively-unimpaired at baseline, and followed for up to eight years. Longitudinal changes in tau PET signal (^18^F-flortaucipir) across 28 bilateral regions, plasma p-tau217, and cognition (Preclinical Alzheimer’s Cognitive Composite [PACC]) were assessed using mixed-effects regression. The correlation between biomarker changes and cognitive decline was estimated using Bayesian joint longitudinal mixed-models.

**Results:** All cognitive outcomes showed strong nonlinear trajectories associated with Aβ-positivity. In Aβ+ participants, the largest effect sizes of longitudinal tau PET accumulation at 36-months were in the inferior temporal gyrus (ITG), fusiform (FUSI), and entorhinal cortex (ERC). Pronounced acceleration over the course of follow-up was observed in the ITG and the middle temporal gyrus (MTG) and the inferior parietal lobule (IPL). Baseline associations with longitudinal change in the PACC were strongest in ERC: correlation (ρ) = −0.55 (−0.63, −0.45), and plasma p-tau217: ρ =−0.47 (−0.56, −0.37). Tau PET changes in frontoparietal regions were strongly correlated with concurrent changes in cognition. Correlations with change in the PACC ranged from −0.65 to −0.61 in the precuneus (PRE), superior parietal lobule (SPL), caudal middle frontal (CMF) and superior frontal gyri. Levels of plasma p-tau217 increased significantly in Aβ+ participants before showing significant deceleration (χ^2^ = 21.7, p < 0.001) and was not associated with concurrent cognitive change, ρ = −0.03 (−0.23 to 0.16).

**Conclusion:** These findings highlight the value of tau PET for both prognostic and real time tracking of disease progression. While plasma p-tau217 may serve as an efficient and scalable biomarker with high prognostic value, region-specific tau PET signals track with concurrent cognitive decline. These results support the inclusion of plasma p-tau217 to guide participant selection and imaging-based tau measures to enhance detection of disease-modifying effects and to refine therapeutic targets in future AD trials.

## Introduction

Developing disease-modifying treatments that result in a durable slowing of clinical decline is a priority for Alzheimer’s disease (AD) research. Disease modification, characterized by an enduring change in the clinical progression of AD by interfering with the underlying pathophysiology of the disease^1^, could be demonstrated by providing evidence of a link between the therapeutic effects on clinical and biomarker endpoints. Reporting these therapeutic effects would demonstrate the biomarker–clinical endpoint association at the trial level (within-trial association between the effect of treatment on biomarker and the effect of treatment on clinical outcome). Additional evidence for the strength of disease modification could be provided by demonstrating that within treatment groups, individual-level biomarker changes are associated with individual changes on the clinical endpoint.^2^ Neither the trial-level nor the individual-level association are commonly reported. Reporting such associations would strengthen the underlying hypothesis that specific biological treatment effects are clinically meaningful.

There have been recent calls for more transparency^3^ and for trials to pre-specify mediation analyses that demonstrate the proportion of the treatment effect on clinical outcomes that operate through specific biomarker pathways.^4^ Although models incorporating multiple outcomes (biomarker and clinical) pose computational challenges, they offer improved estimation and avoid the biases of estimating correlation between changes in multiple measures from separate models.^5,6^ Reporting pre-specified mediation analyses or estimates of joint longitudinal changes in multiple outcomes would be of high value in AD trials research.

Both recently approved treatments for early AD, lecanemab^7^ and donanemab^8^ successfully removed Aβ and also demonstrated a consistent although moderate slowing of clinical decline. During phase III trials, participants on lecanemab showed significantly slower accumulation of tau (using PET) compared with placebo^9^ while no effect of donanemab on the accumulation of tau was observed.^8^ The mechanism by which lecanemab slows clinical decline may be through a combination of Aβ, tau and other pathways including synaptic health, while donanemab, not showing an effect on tau PET, may provide clinical benefit primarily through reduced Aβ toxicity or through effects on tau metabolism not captured by current tau PET methods. The association between therapeutic effects on Aβ (or tau) and clinical endpoints in these trials were not reported, without which the proportion of the treatment effect on the clinical endpoint operating through these biomarker pathways remains unknown. Individual-level associations between biomarkers and clinical measures were also not reported.

Core biomarkers of Aβ and tau are routinely used in AD clinical trials, observational studies,^10–13^ and are considered diagnostic in the research criteria for AD.^14^ However, reports of individual-level concurrent changes of these biomarkers with changes in clinical measures are less common.^15^ This study therefore aimed to estimate the association between concurrent biomarker and cognitive changes in preclinical AD, defined as asymptomatic Aβ pathology. These analyses focus on the longitudinal change in tau PET and plasma p-tau217 and how these changes correlate with concurrent changes in cognition.

## Methods

### Participants

Data were obtained from the Anti-Amyloid Treatment in Asymptomatic Alzheimer’s (A4) and companion study, Longitudinal Evaluation of Amyloid Risk and Neurodegeneration (LEARN).^16^ A4/LEARN were approved by the Institutional Review Boards of all participating institutions. Informed written consent was obtained from all participants at each site. Participants eligible for screening into the A4 Study were age 65-85 and cognitively unimpaired with CDR-Global score 0, MMSE 25-30, and Weschler Memory Scale Revised Logical Memory IIa Delayed Recall score of 6-18. Analyses comparing plasma p-tau217 and tau PET and their relationships with cognitive change were limited to participants with elevated Aβ (A4) with all three outcome measures (N=374). Analyses of individual outcomes included all participants (PACC: N=1707; plasma p-tau217: N=1643, tau PET: N=443) including those without elevated Aβ (LEARN) for reference. Follow-up included both the double-blind phase as well as the open label extension.

### PET Imaging

A4/LEARN ^18^F-florbetapir Aβ PET data were processed using a PET-only pipeline, using an average across six cortical regions from the AAL atlas (anterior cingulate, posterior cingulate, lateral parietal, precuneus, lateral temporal, and medial orbital frontal), and normalized to a whole cerebellum reference region to create standardized uptake value ratios (SUVRs). Aβ-positivity was defined previously using a hybrid quantitative-qualitative approach (SUVR>1.15 or an SUVR between 1.10 and 1.15 with a positive visual read).^17^

For processing 18F-flortaucipir PET data,12 individual-level T1 images were coregistered to the PET data and used to spatially normalize individual-level flortaucipir PET to MNI space. The GTMseg atlas18 was used to extract regional intensities across 28 bilateral tau regions of interest (ROIs): the amygdala, entorhinal cortex, parahippocampal gyrus, fusiform gyrus, banks of the superior temporal sulcus (bankssts), temporal pole, and the inferior/middle/superior temporal gyri; the isthmus cingulate, insula, rostral/caudal anterior cingulate, precuneus, posterior cingulate, supramarginal gyrus, and the inferior/superior parietal lobules; the inferior frontal gyrus (comprising the pars orbitalis, pars triangularis, and pars opercularis), lateral/medial orbitofrontal, rostral/caudal middle frontal cortices, frontal pole and superior frontal gyus; and the cuneus, lingual gyrus, and lateral occipital cortex. Regional intensity values were normalized to a cerebellar grey matter reference region to create standardized uptake value ratios (SUVRs).

### Plasma p-tau217

Plasma p-tau217 was assayed on an analytically validated ECL immunoassay using a MesoScale (MSD) Sector S Imager 600 MM at the CAP-accredited, CLIA-certified Lilly Clinical Diagnostics Laboratory on plasma samples from baseline, week 12, week 72, and week 240 or endpoint from A4, and from week 24, week 240 or endpoint from LEARN.^19^

### Cognition

The four-component Preclinical Alzheimer’s Cognitive Composite Scale (PACC) was used to assess cognition,^20^ comprising the Free and Cued Selective Reminding Test (FCSRT, Free+Total Score, range 0-96), Logical Memory IIa delayed recall (range 0-25), the Digit-Symbol Substitution Test, and the Mini Mental Status Examination (MMSE) (range 0-30). Three versions of the PACC components were alternated across visits to minimize practice effects.^16^ To calculate the PACC, each component was standardized to their respective baseline mean and standard deviation among Aβ– participants (N=538). These four z-scores were then summed and z-scored again to the baseline mean and standard deviation of the Aβ– participants. Each PACC component was also examined.

### Statistical Analysis

Each of the outcomes (tau PET, p-tau217, and the PACC) were first modeled separately and summarized. Then all three outcomes were modeled jointly for estimates of the correlation between outcomes. In the first set of analyses, repeated measures of ^18^F-flortaucipir PET SUVR were modeled using mixed-effects regression assuming participant-specific random intercepts and slopes. Natural cubic splines were used to capture any departure from linearity in the accumulation of tau over time. The significance of nonlinear change over time was tested by comparing a model assuming a linear time term and a model incorporating spline expansion terms for time using a likelihood ratio test. The number of spline knots was chosen to minimize the Akaike Information Criterion (AIC). Tau PET models that focused on accumulation rates in Aβ+ included age, sex, treatment (solanezumab vs placebo, assignment at randomization) and two spline expansion terms for years since baseline scan. In models that compared trajectories of Aβ+ and Aβ– participants, treatment was not included in the model, given that treatment was not associated with changes in the PACC or tau PET.^16^ The interaction between Aβ status and the spline time terms was included. Tau ROIs were compared on the estimates of change at 36 months, expressed as effect sizes, calculated as change at 36 months divided by the model residual standard error, and 95% confidence intervals. Longitudinal plasma p-tau217 was modeled similarly to tau PET.

To assess acceleration of tau accumulation over the course of follow-up, numerical differentiation using finite differences was used to approximate the first derivative of the regression curves of longitudinal tau PET SUVR. These curves were then used to estimate a positional variance diagram to rank the ROIs in terms of their instantaneous accumulation rate at 36 months. The positional variance diagram shows the proportion of 1000 bootstrap samples in which each ROI appears in a particular position in the central ranking.

In the second set of analyses, repeated measures of cognitive outcomes were modeled similarly to the tau PET or plasma p-tau217 outcomes. These models included age, sex, years of education, test version, treatment, and spline expansion terms for years since baseline. Similar to the tau PET analyses, for models comparing Aβ+ and Aβ– trajectories, treatment was omitted while the interaction between Aβ status and the spline terms was included in the model. PACC models included three spline expansion terms for time since baseline.

In the third set of analyses, Bayesian joint longitudinal mixed models^21^ were used to estimate the associations between the participant-specific random intercepts and slopes of tau PET, plasma p-tau217, and the PACC. Model estimation was performed via Markov Chain Monte Carlo using four chains and 3000 iterations (1500 warm-up). R-hat values and trace plots were inspected for convergence. Analysis of residuals and posterior predictive checks were evaluated for model fit.^22^ Because all outcomes were skewed due to small numbers of individuals with high tau PET SUVRs, plasma p-tau217 levels or low PACC scores, all outcomes were Box-Cox transformed^23^ and then centered and scaled to their respective mean and standard deviation. Values were back-transformed to the original scales for depiction in figures.

Two sensitivity analyses were performed. The first was an adjustment of tau PET outcomes for the interaction between global tau burden at baseline and the spline terms for time. Global tau burden was taken to be the unweighted average of the 28 tau PET ROIs at baseline. This was done to assess whether the magnitude of the associations between regional tau and the PACC differed within a given level of overall tau severity. The second sensitivity analysis was an adjustment of p-tau217 for the interaction between global Aβ burden (PET) at baseline and the spline terms for time.

Finally, participant-specific random intercept and slope estimates from plasma p-tau217, PACC and region-specific tau PET (ITG, IPL, PRE, and CMF) mixed-effects models were submitted to Gaussian mixture models to explore clustering among participants. The optimal number of clusters was selected according to the Bayesian Information Criterion (BIC).

## Results

### Cohort Characteristics

A total of 1707 participants (1169 Aβ+, 538 Aβ–) were included in the analyses. Participants were 71.5 years old on average (range: 65-85), 60% female, had an average of 16.6 years of education, an average baseline MMSE score of 28.8, and 48% carried at least one APOE ε4 allele. The tau PET substudy included a total of 443 participants (388 Aβ+, 55 Aβ–). Tau PET substudy participants were 71.8 years old on average (range: 65-85), 58% female, had an average of 16.2 years of education, an average baseline MMSE score of 28.7, and 53% carried at least one APOE ε4 allele. Single outcome models included all available data (plasma p-tau217: N=1643; cognitive models: N=1707).

### Regional Tau PET Accumulation Rates

Participants in the tau PET substudy had between 1–5 scans during the course of the study: N=49 had one scan, 56 had two, 92 had three, 135 had four and 111 had five scans. Median follow-up time was 4.8 years (range: 0-8.1 years).

Longitudinal trajectories of Aβ+ vs Aβ– participants for six select ROIs are shown in Figure 1A. Twenty-two of the 28 ROIs showed significant increases in tau PET SUVR 36 months after baseline in the Aβ+ participants (Figure 1B). In Aβ+ participants, the fastest accumulating regions were ITG: 36-month change from baseline effect size [ES] = 1.12 (0.93, 1.31) and FUSI: ES = 1.04 (0.86, 1.22).

**Figure 1.**
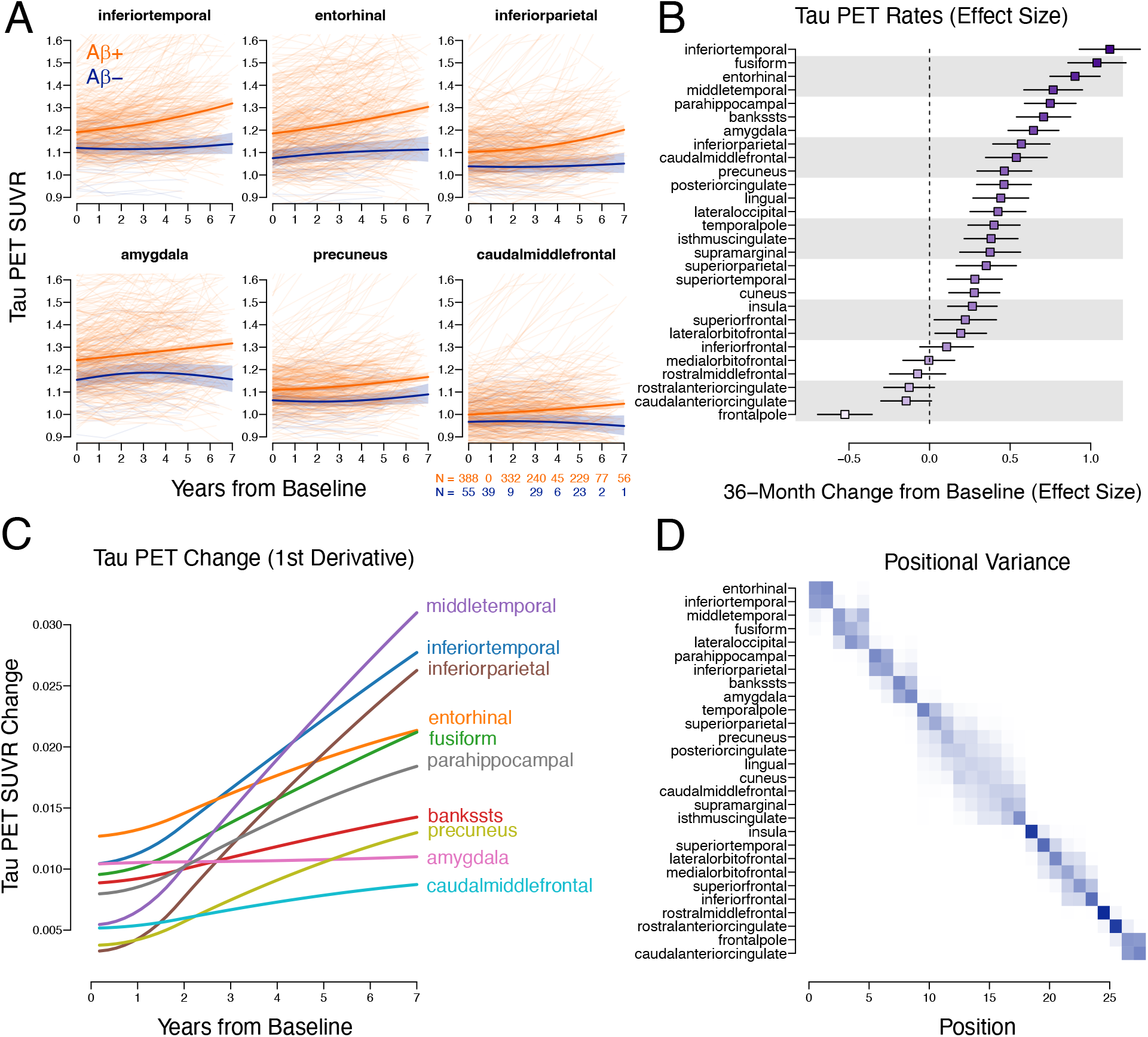
Longitudinal tau PET trajectories. In panel A, longitudinal curves of tau PET for Aβ+ (orange) and Aβ– participants (blue) are shown over seven years of follow-up. Spaghetti plots of individual observed SUVRs are shown, as well as population curves with 95% confidence intervals (shaded areas). Sample sizes at each year of follow-up are shown in the bottom right. In panel B, estimates of 36-month SUVR change from baseline, expressed as an effect size, with 95% confidence intervals are shown for all 28 ROIs. In panel C, flortaucipir PET change curves, estimated by the first derivative of the SUVR curves in the Aβ+ participants are shown over seven years of follow-up. In panel D, all ROIs are ranked on their 36-month estimated rate of change based on the curves shown in panel C. The strength of the shading represents the proportion of bootstrap samples resulting in the indicated column rank. More shaded ranks in a row indicate more positional variation.

Significant acceleration of accumulation was observed in 15 of the 28 ROIs, including ITG (χ^2^ = 11.8, p = 0.001), MTG (χ^2^ = 34.4, p < 0.001), and IPL (χ^2^ = 32.9, p < 0.001), but not ERC (χ^2^ = 2.0, p = 0.15) or the amygdala (χ^2^ = 0.0, p = 0.93). Tau PET change (rather than SUVR), estimated using the first derivative of the population SUVR curves of the Aβ+ participants is shown in Figure 1C. These curves depict the acceleration of accumulation over time, with ITG, MTG, and IPL showing pronounced acceleration over the seven years of follow-up, while ERC showed the highest estimated instantaneous rate of accumulation at baseline. ROI ranks, in terms of tau PET 36-month instantaneous rate of change are shown in the positional variance diagram in Figure 1D. ERC, ITG, MTG, and FUSI rank highest for the instantaneous rate of change at 36 months after baseline.

### Plasma p-tau217

Longitudinal trajectories of Aβ+ vs Aβ– participants are shown in Figure 2A. The effect size for 36-month change in p-tau217 in Aβ+ participants was 1.19 (1.06, 1.33), Figure 2A). Plasma p-tau217 also showed eventual significant deceleration in Aβ+ participants (χ^2^ = 21.7, p < 0.001).

**Figure 2.**
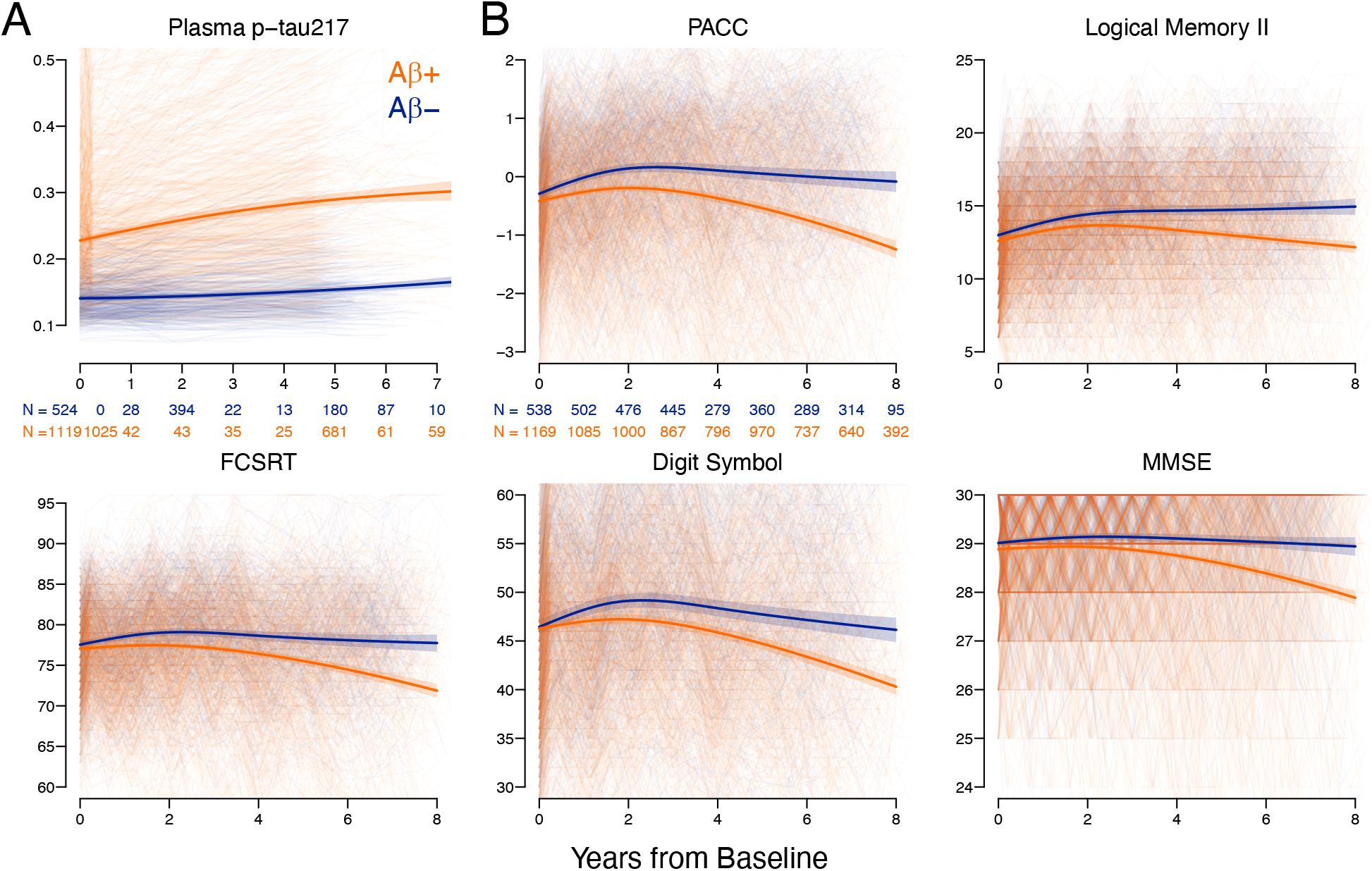
Longitudinal plasma p-tau217 and cognitive trajectories. Longitudinal curves of plasma p-tau217, the PACC, and the individual PACC components, for Aβ+ (orange) and Aβ– participants (blue) are shown over eight years of follow-up. Spaghetti plots of individual observed p-tau217 or cognitive scores are shown, as well as population curves with 95% confidence intervals (shaded areas). Sample sizes at each year of follow-up are shown in the plot in the upper left plots (p-tau217, PACC).

### Cognitive Changes

All cognitive trajectories were strongly associated with Aβ positivity (Figure 2B). The strongest trajectory differences between Aβ+ and Aβ– over the entire course of follow-up was on the PACC (χ^2^ = 86.7, p < 0.001, ΔAIC =−81), followed by Digit Symbol Substitution (χ^2^ = 78.5, p < 0.001, ΔAIC =−73), Logical Memory II (χ^2^ = 65.4, p < 0.001, ΔAIC =−59), MMSE (χ^2^ = 62.9, p < 0.001, ΔAIC =−57), and FCSRT96 (χ^2^ = 59.2, p < 0.001, ΔAIC =−53).

### Associations among PACC change, Tau PET and Plasma p-tau217

The correlation (ρ) of longitudinal changes in the PACC with baseline tau PET ROIs and baseline plasma p-tau217 in Aβ+ participants are shown in Figure 3A. The strongest correlations among the baseline biomarker measures with PACC change were in ERC: ρ=−0.55 (−0.63,-0.45) and plasma p-tau217: ρ=−0.47 (−0.56,-0.37).

**Figure 3.**
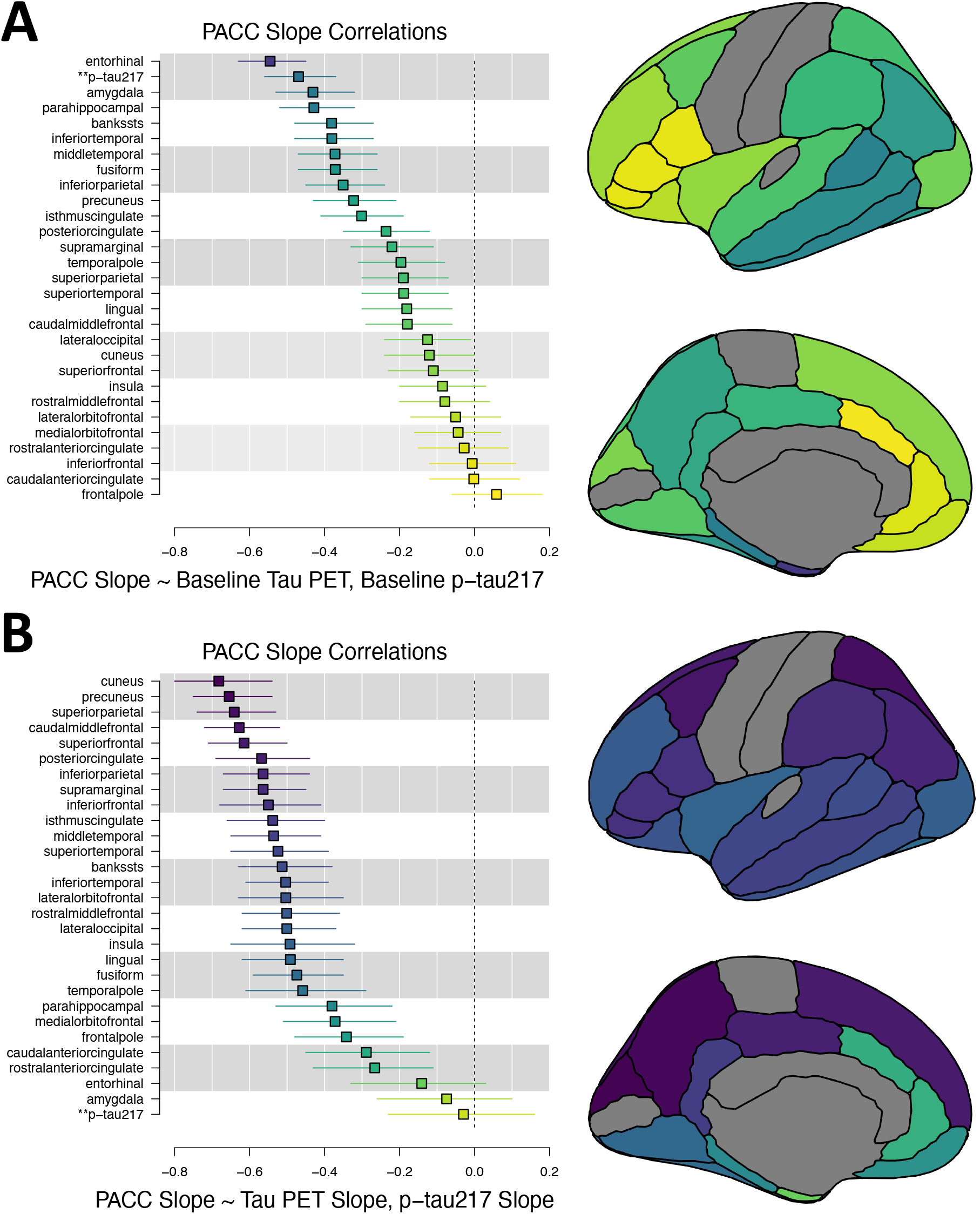
PACC slope correlation with tau PET and plasma p-tau217 in Aβ+ participants. In panel A, the correlation between the individual-specific slopes for change in the PACC and individual-specific <u>intercepts</u> for both the tau PET ROIs and p-tau217 are plotted along with 95% confidence intervals. Plasma p-tau217 is shown with leading **. Tau PET ROIs, colored according to correlation estimates are shown on the right. In panel B, the correlation between the individual-specific slopes for change in the PACC and individual-specific <u>slopes</u> for both the tau PET ROIs and p-tau217 are shown.

The correlation of concurrent longitudinal changes in the PACC with longitudinal changes in tau PET ROIs and plasma p-tau217 in Aβ+ participants are shown in Figure 3B. The strongest correlations with PACC change were in frontoparietal regions including change in PRE: ρ=−0.65 (−0.75,-0.54), SPL: ρ=−0.64 (−0.74, −0.53), CMF: ρ=−0.63 (−0.72, −0.52), superior frontal gyrus: ρ=−0.61 (−0.71, −0.50) and in the cuneus: ρ= −0.68 (−0.8, −0.54). Plasma p-tau217 change was not associated with PACC change: ρ=−0.03 (−0.23, 0.16).

The correlation of longitudinal changes in tau PET with baseline plasma p-tau217 are shown in Figure S1. Baseline p-tau217 showed widespread correlations with subsequent longitudinal tau PET change in temporal, but also parietal and frontal regions.

The correlation of longitudinal changes in tau PET with longitudinal changes in plasma p-tau217 are shown in Figure S2. Plasma p-tau217 change also showed widespread correlations with longitudinal tau PET change.

In the first of two sensitivity analyses, after adjusting the tau PET outcomes for the interaction between global tau level at baseline and the spline terms for time, the correlation between the participant-specific intercepts for tau PET and the participant-specific slopes for the PACC were greatly reduced, as expected (corresponding to the main analysis in Figure 3A). On average, the magnitude of the correlation was reduced by 0.26, with many ROIs becoming nonsignificant. In contrast, the adjustment for global tau level had little effect on the correlation between participant-specific slopes for tau PET and slopes for the PACC (corresponding to the main analysis in Figure 3B), where the average reduction in the magnitude of correlation was < 0.01.

In the second sensitivity analysis, the adjustment of the p-tau217 model for the interaction between global Aβ PET and the spline terms for time reduced the magnitude of correlation between participant-specific intercepts of p-tau217 and the slopes of the PACC by 0.06 (corresponding to the main analysis in Figure 3A). Plasma p-tau217 remained uncorrelated with PACC change after the adjustment for global Aβ PET.

Individual-specific slope estimates of change in the PACC are plotted against both baseline and slopes of tau PET in Figure 4A, colored by class, estimated by Gaussian mixture models. Slope estimates of tau PET are plotted against baseline and slopes of p-tau217 in Figure 4B. And slopes of the PACC are plotted against baseline and slopes of plasma p-tau217 in Figure 4C. Three classes were identified based on patterns of the baseline and slopes of four tau PET ROIs (ITG, IPL, PRE, and CMF), plasma p-tau217, and the PACC. Class 3 had generally higher Aβ SUVRs, p-tau217, ERC tau, and worse PACC scores at baseline on average (Table).

**Figure 4.**
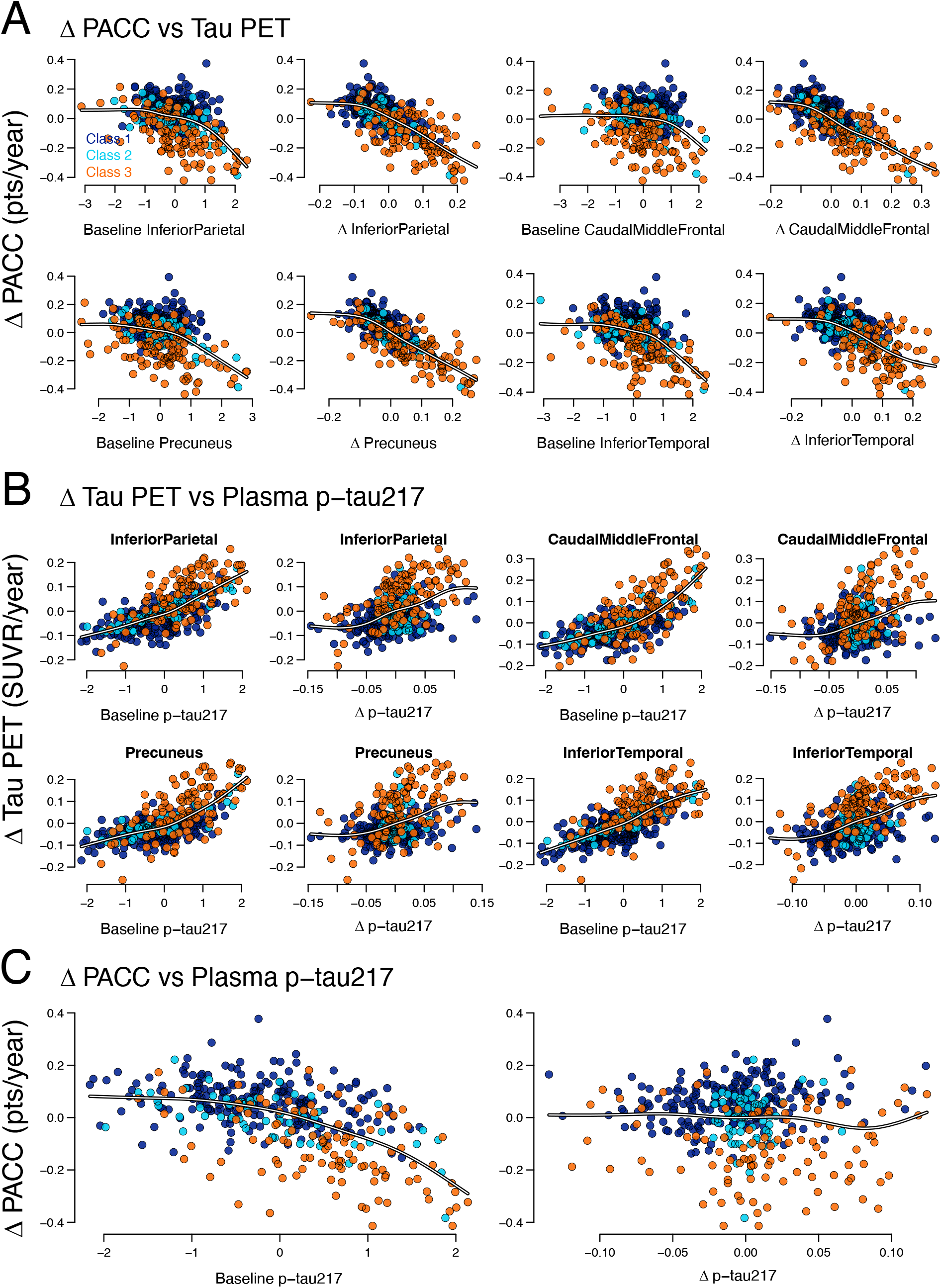
PACC, tau PET, and plasma p-tau217 correlations and class membership in Aβ+ participants. In panel A, individual-specific slopes for the PACC are plotted against individual-specific intercepts and slopes of four select tau PET regions. Class membership is shown with class 1 in dark blue, class 2 in turquoise, and class 3 in orange. In panel B, individual-specific slopes for the four tau PET regions are plotted against individual-specific intercepts and slopes of plasma p-tau217. In panel C, individual-specific slopes for the PACC are plotted against individual-specific intercepts and slopes of plasma p-tau217.

**Table.** Baseline Class Characteristics.

|  | Class 1 | Class 2 | Class 3 |
| --- | --- | --- | --- |
| N | 209 (55.9%) | 61 (16.3%) | 104 (27.8%) |
| Age | 71.7 (4.6) | 72.4 (4.7) | 72.7 (5.2) |
| Sex, F | 55 (40.0%) | 40 (65.6) | 65 (62.5) |
| Education | 16.2 (2.7) | 16.1 (3.8) | 16.2 (2.5) |
| TX, active | 115 (55.0%) | 25 (41.0%) | 45 (43.3%) |
| Aβ SUVR | 1.28 (0.15) | 1.30 (0.19) | 1.39 (0.18) |
| p-tau217 | 0.24 (0.12) | 0.26 (0.14) | 0.35 (0.18) |
| ERC SUVR | 1.16 (0.12) | 1.23 (0.16) | 1.28 (0.21) |
| PACC | -0.81 (1.1) | -0.97 (1.1) | -1.18 (1.1) |
| MMSE | 28.7 (1.4) | 28.6 (1.2) | 28.4 (1.4) |

## Discussion

The potential of two biomarkers—tau PET and plasma p-tau217—as candidates to establish disease modification in preclinical AD was examined. These findings suggest that both biomarkers demonstrate strong predictive value for cognitive decline, while only tau PET uptake, particularly in frontoparietal regions showed strong associations with concurrent changes in cognition.

Previous estimation of tau PET rates of change in preclinical AD have been limited by short follow-up times and small numbers of serial scans within individuals. Due to the extensive follow-up in A4, with more than half of the tau PET cohort having four or five scans, acceleration in tau accumulation was detectable. Previously reported in two large longitudinal cohorts^24^ and now replicated here in A4, the inferior temporal gyrus is the fastest accumulating region for tau in preclinical AD. The inferior temporal gyrus is also a site of Aβ deposition that has been shown to interact with local tau to facilitate widespread tau propagation to neocortical regions.^25^ Other rapidly accumulating regions included the entorhinal cortex and fusiform gyrus. Regions with the most pronounced acceleration included the inferior and middle temporal gyri, inferior parietal lobule, and the lateral occipital cortex.

The associations observed between baseline levels of tau PET and plasma p-tau217 with subsequent cognitive decline suggest that both biomarkers could serve as valuable tools for early risk stratification for recruitment into AD trials. Plasma p-tau217 may become abnormal earlier in the disease course than tau PET^14,26^ presumably reflecting Aβ-related tau phosphorylation upstream of tau tangle formation. Plasma p-tau217’s early changes and predictive ability provide an early, systemic indicator of tau pathology. Plasma p-tau217 has been shown to be strongly associated with longitudinal changes in Aβ PET, within Aβ– individuals.^27^ Plasma p-tau217 has also been shown to be more strongly correlated with Aβ PET than tau PET in early stages of AD and also to mediate the association between Aβ PET and tau PET.^28^ Because plasma p-tau217 becomes abnormal near the time Aβ PET becomes abnormal, plasma p-tau217 has been thought to be more closely related to Aβ pathology. However, given the association observed here between p-tau217 and tau PET (Figure S1, S2, 4B) and also the ability of plasma p-tau217 to predict future cognitive changes beyond that of Aβ PET,^12^ it is likely that plasma p-tau217 is a more direct measure of tau pathology.

Baseline plasma p-tau217’s predictive capacity for cognitive change was similar to early tau PET regions. Among tau PET regions, the entorhinal cortex stands out as the only region whose point estimate of tau burden to predict cognitive changes exceeded that of plasma p-tau217 (Figure 3A). This finding aligns with the entorhinal cortex’s role as a hub in early AD pathology.^29^ Notably, while the entorhinal cortex and the amygdala were among the regions most highly associated with future cognitive decline, they were the only two tau PET regions not associated with concurrent cognitive decline, similar to plasma p-tau217, possibly indicating a plateauing of accumulation in participants with the most pronounced cognitive decline.

The observed association between changes in tau PET levels—particularly in frontal and parietal regions (Figure 3B)—and concurrent changes in cognition indicates that tau PET may serve as a valuable marker for tracking disease progression in real time. In clinical trials, tau PET’s dynamic association with cognition could provide an early, intermediate measure of treatment effects, making it particularly useful for evaluating intervention efficacy and disease modification. The acceleration of accumulation in many tau PET regions aligns with the acceleration of cognitive decline seen on the PACC. The deceleration of plasma p-tau217 (figure 2A) likely mitigates its association with both changes on the PACC and changes in tau PET.

This study has several limitations. The ^18^F-flortaucipir ligand has known off-target binding issues occurring in the hippocampus, precluding evaluation of hippocampal tau.^30^ The participants in A4 and LEARN were volunteers, meeting screening criteria including requirements for general good health and likely do not represent the general population. The A4/LEARN cohorts also do not represent the racial and ethnic diversity of the general population, which may influence the relationships between tau biomarkers and cognition.^12^

Selecting specific brain regions to represent tau pathology in AD should be based on specific research goals. Studies concerning rapid tau accumulation in preclinical AD may be optimized by focusing on the inferior temporal gyrus, while prediction of future cognitive decline may be better served by tau levels in the entorhinal cortex or plasma p-tau217. For tracking real time cognitive change, frontoparietal tau PET may be optimal. These regions may be especially valuable for evaluating disease-modifying effects in clinical trials. Although these findings underscore the potential of tau PET as a marker of disease modification, it remains unknown whether it can reliably forecast treatment-related cognitive benefits. Confirming these results in broader, more diverse populations and across various interventions, with long-term follow-up, will be important for generalizing these results.

## Data Availability

All data produced are available online at
https://www.synapse.org/Synapse:syn61250768/wiki/628717

## Acknowledgements

The authors would like to thank the entire A4 and LEARN Study Teams, including all site principal investigators, and their dedicated site staff. Special gratitude to the A4 and LEARN participants and their study partners, without whom these studies would not be possible.

## Ethical Considerations

Approval from an institutional review board or ethics committee was obtained at each of the sites. All participants and their study partners provided written informed consent prior to data collection, which included consent for data sharing.

