## Supplemental Material for "Concurrent longitudinal changes in plasma p-tau217, tau PET, and cognition in preclinical Alzheimer’s disease"

Figure S1. Tau slope correlation with baseline plasma p-tau217 in Aβ+ participants

The correlation between the random slopes for change in tau PET ROIs and random intercepts for plasma p-tau217 are plotted along with 95% confidence intervals.

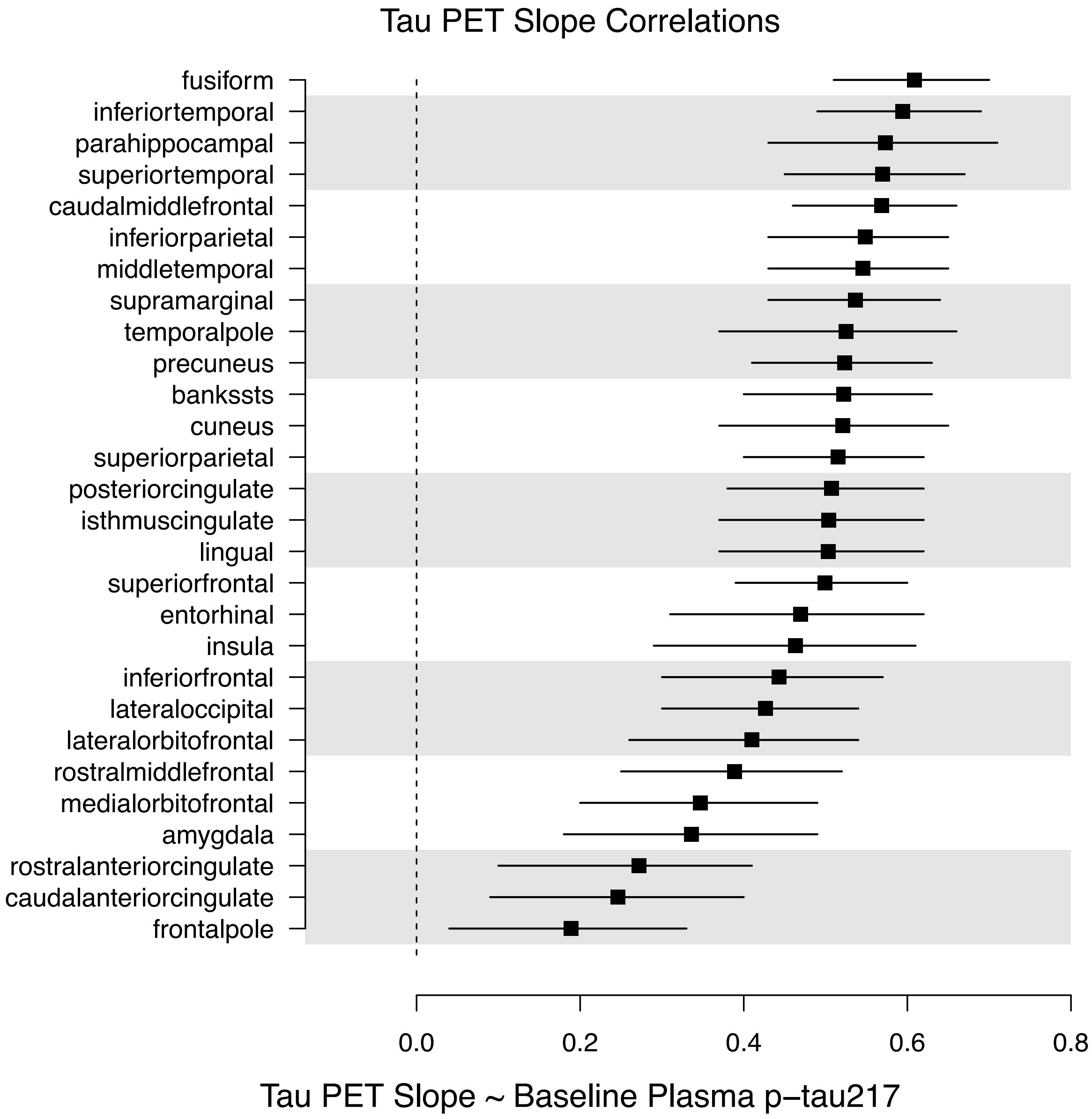

Figure S2. Tau slope correlation with plasma p-tau217 slope in Aβ+ participants

The correlation between the random slopes for change in tau PET ROIs and random slopes for change in plasma p-tau217 are plotted along with 95% confidence intervals.

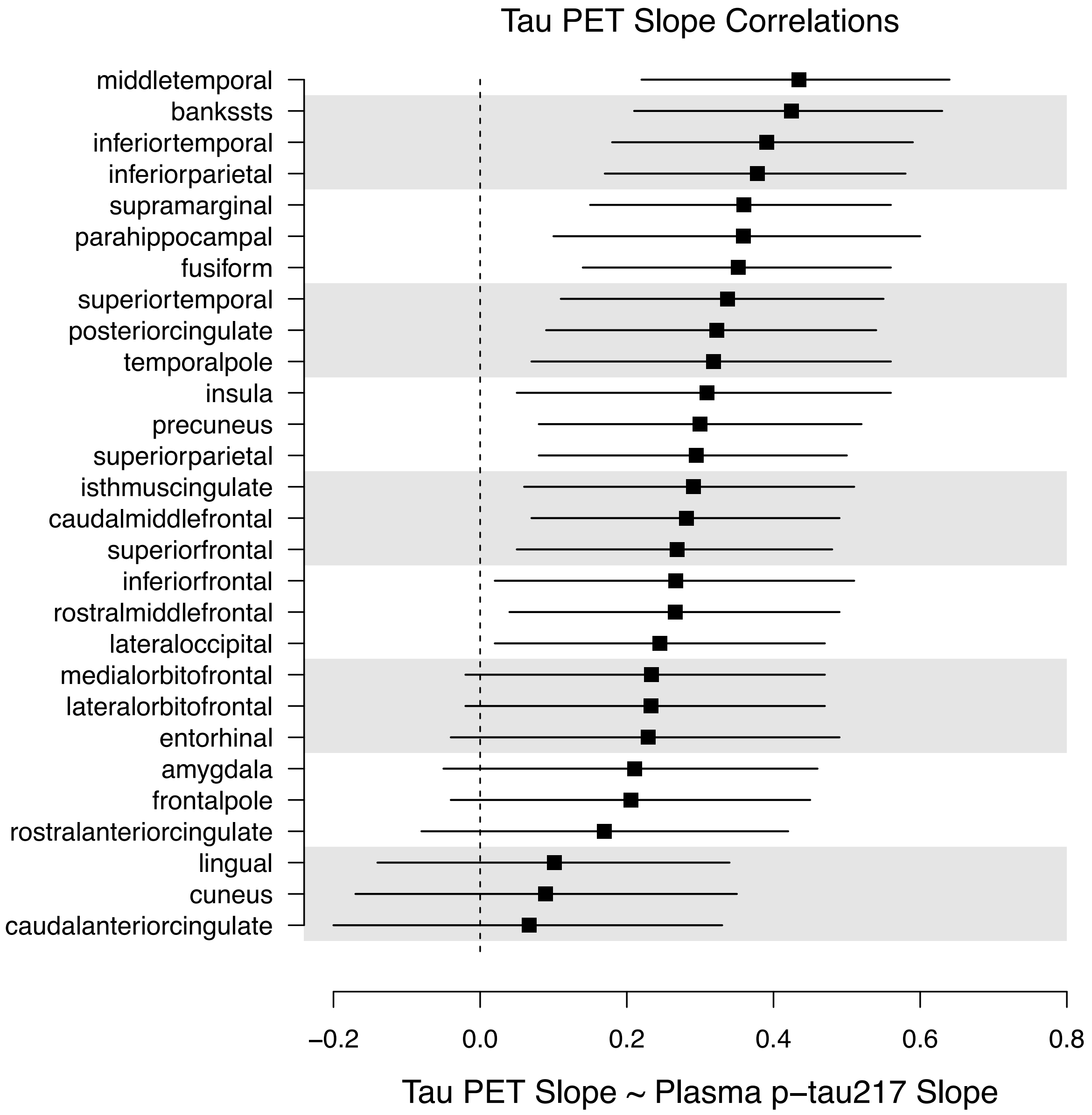
